# Preconception Genetic Carrier Screening for Miscarriage Risk Assessment: A Bioinformatic Approach to Identifying Candidate Lethal Genes and Variants

**DOI:** 10.1101/2023.05.25.23290518

**Authors:** Mona Aminbeidokhti, Jia-Hua Qu, Shweta Belur, Hakan Cakmak, Eleni Jaswa, Ruth B. Lathi, Marina Sirota, Michael P. Snyder, Svetlana A. Yatsenko, Aleksandar Rajkovic

## Abstract

**Purpose:** Miscarriage, due to genetically heterogeneous etiology, is a common outcome of pregnancy. Preconception genetic carrier screening (PGCS) identifies at-risk partners for newborn genetic disorders; however, PGCS panels currently lack miscarriage-related genes. Here we assessed the theoretical impact of known and candidate genes on prenatal lethality and the PGCS among diverse populations.

**Methods:** Human exome sequencing and mouse gene function databases were analyzed to define genes essential for human fetal survival (lethal genes), identify variants that are absent in a homozygous state in healthy human population, and to estimate carrier rates for known and candidate lethal genes.

**Results:** Among 138 genes, potential lethal variants are present in the general population with a frequency of 0.5% or greater. Preconception screening for these 138 genes would identify from 4.6% (Finnish population) to 39.8% (East Asian population) of couples that are at-risk for miscarriage, explaining a cause for pregnancy loss for ∼1.1-10% of conceptions affected by biallelic lethal variants.

**Conclusion:** This study identified a set of genes and variants potentially associated with lethality across different ethnic backgrounds. The diversity of these genes amongst the various ethnic groups highlights the importance of designing a pan-ethnic PGCS panel comprising miscarriage-related genes.

## INTRODUCTION

Human reproduction is inefficient, with ∼70% of all conceptions not progressing to live birth across different developmental stages.^1,2^ Reproductive failures commonly occur in the early stages post-conception, with ∼10-30% of embryos being developmentally arrested before implantation.^1-3^ An additional 30% of losses occur after implantation within 3-4 weeks of gestation before the pregnancy is even recognized.^1^ Finally, 10-20% of clinically recognized pregnancies are lost in the first trimester.^1^

Fetal chromosomal abnormalities associate with 50% of first-trimester miscarriages, while the etiology of pregnancy loss in the remaining 50% of cases with normal karyotype and chromosomal microarray analysis remains unexplained.^4^ Recent exome sequencing studies have begun to uncover deleterious variants in specific genes that cause euploid pregnancy loss.^5–10^ However, the spectrum of genes associated with human lethality and pregnancy loss remains understudied.

Identifying genes linked to pregnancy losses in humans is a challenging task, mainly due to the lack of large-scale studies that assess pregnancy losses from fertilization to clinical presentation and the appropriate phenotyping of such losses. Moreover, damaging variants in some critical genes may affect early stages of development and not be associated with any particular phenotype archived in the Online Mendelian Inheritance in Man (OMIM) database.

Out of the ∼20,000 human genes, only a quarter is associated with any known phenotype, the majority of which is observed postnatally.^11^ Based on the current entries in OMIM, it has been estimated that only 624 (3.7%) of human genes are associated with lethal phenotypes that may result in fetal or perinatal mortality.^11^ The actual set of human genes essential to maintain fetal viability may be much larger as 39.2% of mouse genes are associated with lethality through knockouts.^11,12^ Previous studies have guesstimated that ∼3,400 of human genes are essential for embryonic and fetal survival.^11^ The potential contribution of candidate genes to human lethal phenotypes and pregnancy losses is unknown.

The objective of this study was to use existing human exome datasets and discern genes that could be potentially added to preconception genetic carrier screening (PGCS) to identify couples at risk for pregnancy loss. We also used a current database of human exomes to evaluate the frequency of pathogenic, likely pathogenic, and putative loss-of-function variants in targeted genes.^13,14^ We calculated the distribution of such variants across different ancestral populations and the probability that such variants contribute to human pregnancy losses. We identified 138 known and candidate lethal genes that have a high-frequency rate of ≥0.5% (1/200 individuals) of potentially damaging variants and are present among different ethnic groups. These 138 genes may significantly contribute to pregnancy losses, and may be included as part of PGCS, which will allow preimplantation genetic testing to avoid pregnancy loss for at-risk couples. Future studies will be needed to determine the validity of our conclusions.

## MATERIALS AND METHODS

### Selection of lethal candidate genes and variant filtration

In this study, we investigated genes related to human lethality, including “known lethal genes” and “candidate lethal genes”. Among nuclear genes (16,742) deposited in OMIM, 6,406 genes had associated human phenotype. Perinatal lethal human phenotypes were documented for 624 genes,^11^ including 120 genes with autosomal dominant, 17 genes with an X-linked, and 487 genes with autosomal recessive inheritance. For the purpose of this study, we focused on 487 genes with autosomal recessive inheritance, defined as “known lethal genes” (Figure 1). ClinVar curated pathogenic (P), and likely pathogenic (LP) variants were downloaded from the Genome Aggregation Database (gnomAD) v2.1.1.^15^ In addition, loss-of-function (LoF) variants including nonsense, frameshift, insertions/deletions, and splice site alterations that disrupt the reading frame of protein-coding genes, were obtained from gnomAD and included in the analysis. For each variant, the extracted parameters contained allele frequencies for the entire population acquired from 125,748 exome sequences, as well as for available ethnic groups: African/African American, Ashkenazi Jewish, East Asian, Finnish, Latino/Admixed American, Non-Finnish European, and South Asian (Supplemental data, Table S1).

**Figure 1.**
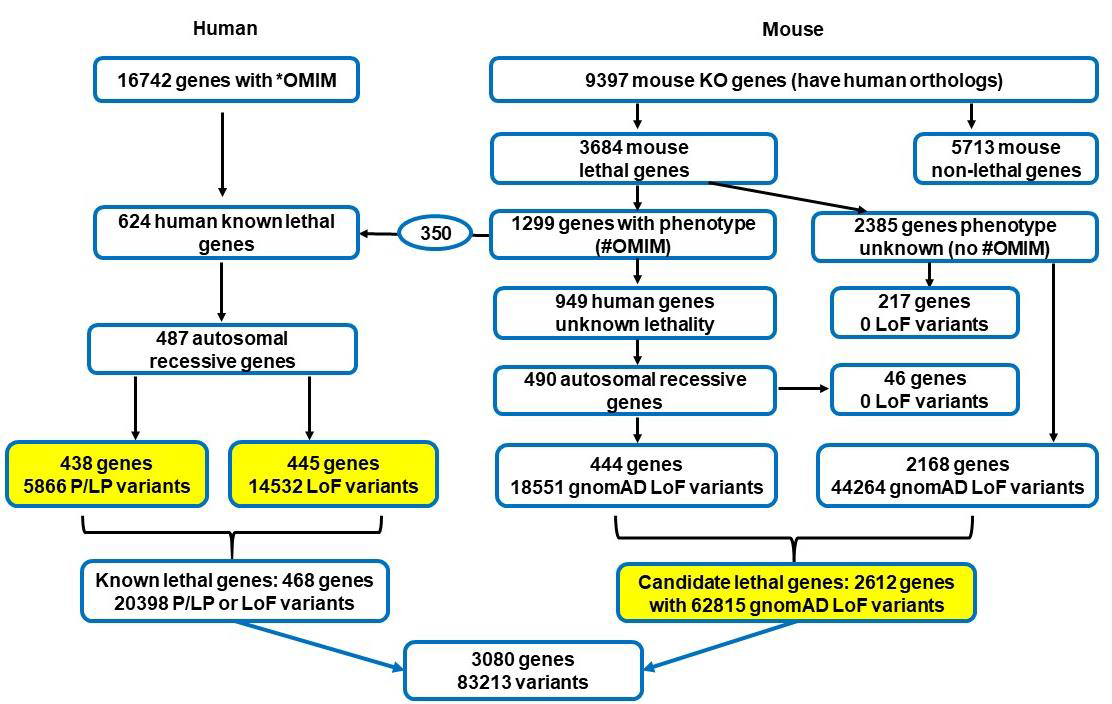
A bioinformatic pipeline for the identification of known and candidate genes related to human lethality. OMIM database was screened to filter genes associated with human lethality (624 human known lethal genes). Genes with an autosomal recessive inheritance were selected for further analysis of the corresponding pathogenic/likely pathogenic (P/LP) and loss-of-function (LoF) variants not classified as P/LP. A set of candidate genes (potentially lethal in humans) has been derived from a list of genes with lethal mouse knock-out (KO) phenotypes. In the OMIM database, human phenotype descriptions were available for 1,299/3,684 genes, including 350 genes currently known to be associated with prenatal lethality in humans. Those 350 genes were already included in the set of “known lethal genes”. The remaining 949 genes had human phenotype descriptions in OMIM but variants in these genes have not reported as lethal in OMIM yet. Out of 949, “490 genes with a recessive inheritance pattern were selected for further analysis. For the remaining 2,385/3,684 candidate genes, no OMIM entry was available, and these genes had no known associated clinical phenotype or mode of inheritance. Overall, 2,875 (490+2,385) candidate genes were subjected to a population-based analysis and search for LoF variants documented in the gnomAD database, yielding findings on 2,612 genes. The total number of genes and their corresponding P/LP and LoF variants are shown at the bottom. In yellow, three datasets subjected to analysis.

We also generated a human “candidate lethal gene” list, derived from a set of 9,397 genes with mouse knock-out phenotypes extracted from the Mouse Genome Informatics and International Mouse Phenotyping Consortium databases as previously described.^11^ Knockouts for 3,684 genes, which had human orthologues, induced pre-weaning lethality in mice. A series of filters (supplemental data) was applied to extract LoF variants observed in a heterozygous but not in a homozygous state in 125,748 exomes of healthy adults (Figure 1). We assumed that these variants are functional, fully penetrant, and are inherited in an autosomal recessive fashion in humans as in mouse models. We excluded structural, missense, or other types of variants due to the uncertainty of their functional significance in candidate genes.

### Population risk probability metrics

To estimate population risk probabilities, calculations were done on three sets of variants: P/LP variants in the “known lethal genes”, LoF variants in the “known lethal genes”, and LoF variants in the “candidate lethal genes”. The following metrics were computed as previously described^14^: variant carrier rate (VCR) – frequency of a variant in the general population; gene carrier rate (GCR) – a cumulative frequency of all P/LP or LoF variants in a given gene, and at-risk couples rate (ACR) – the proportion of couples where both partners carry a likely lethal variant in the same gene (supplemental data). Calculations were also performed for each ancestry.

### Gene ontology annotations of known and candidate lethal genes

The top genes with GCR≥0.005 were subjected to ontology analyses using the PANTHER (Protein Analysis Through Evolutionary Relationships) Classification System version-17 (http://pantherdb.org). Sixty-one “known lethal genes” and 77 “candidate lethal genes” were annotated according to the Gene Ontology Biological Process, Molecular Function, Cellular Component enrichment, Protein class and implicated Pathways.

## RESULTS

### Lethal gene variants in the general population

In 125,748 exomes of healthy individuals, among 487 “known lethal genes”, P/LP and LoF variants were detected in 438 and 445 genes, respectively. Overall, 468 genes had at least one P/LP and/or LoF variants, while for 19 genes no P/LP or LoF were recorded (Figure 1). In gnomAD 62815 LoF variants were annotated for 2,612 genes in the general human population and included in the “candidate lethal genes” analysis. Calculations were made for three sets: 5,866 P/LP variants (all types classified as P/LP by ClinVar) in 438 “known lethal genes”; 14,532 LoF variants (not classified as P/LP by ClinVar) in 445 “known lethal genes”; and 62,815 LoF variants in 2612 “candidate lethal genes” (Figure 1). Among the “known lethal genes”, 94/5,866 (1.6%) of P/LP and 54/14,532 (0.37%) of LoF variants were seen with a frequency of **≥**0.5% or 1/200 individuals in the general population (supplementary Figure S1A,B). In “candidate lethal genes”, 519/62,815 (0.83%) of LoF variants had VCR**≥**0.5%. The remaining LoF variants were rare with VCR<0.5% in the general population (Supplementary Figure S1C).

### Gene carrier rates for P/LP and LoF variants in the “known lethal genes”

A cumulative frequency of variants in a given gene (GCR) was calculated for P/LP variants in “known lethal genes”, LoF variants in “known lethal genes”, and LoF variants in “candidate lethal genes”. In the general population, GCR**≥**0.5% was identified for 9/438 known genes with P/LP variants (*ABCC6* [MIM *603234]*, DHCR7* [MIM *602858]*, F7* [MIM *613878]*, HBB* [MIM *141900]*, KIAA0586* [MIM *610178]*, PKHD1* [MIM *606702]*, PMM2* [MIM *601785]*, SBDS* [MIM *607444]*, SPINK5* [MIM *605010]), 7/445 known genes with LoF variants (*ABCC6* [MIM *603234]*, CEP290* [MIM *610142]*, F7* [MIM *613878]*, KIAA1109* [MIM *611565]*, NPC2* [MIM *601015]*, PKHD1* [MIM *606702]*, SPEG* [MIM *615950]), and 77/2,612 candidate genes with LoF variants (Figure 2, Table S2).

**Figure 2.**
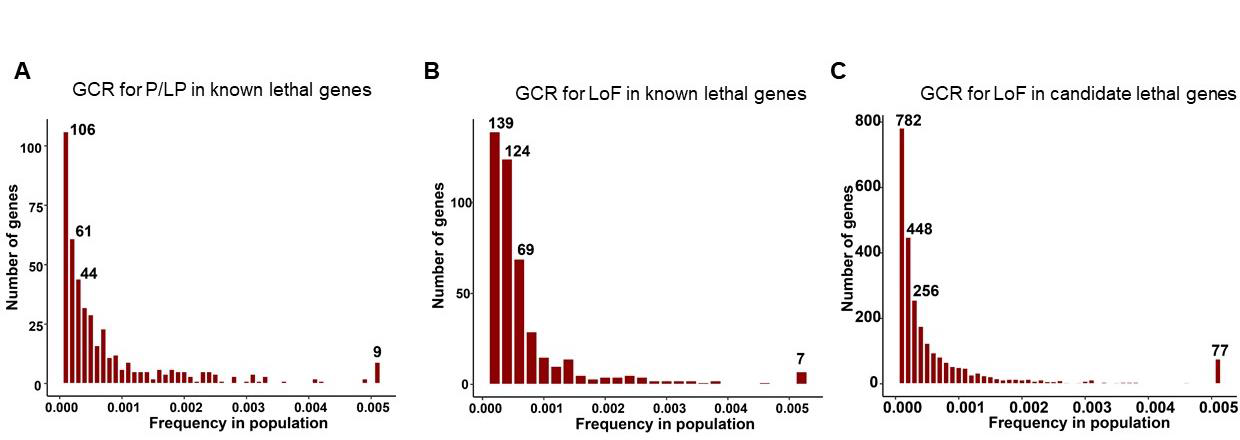
Gene carrier frequency with qualified variants in the general population. (**A**) GCR of P/LP variants in the known lethal genes. Note, 0.5% of individuals in general population (0.005 frequency) carry a P/LP variant in one out of 9 known lethal genes. (**B**) GCR of LoF variants in the known lethal genes. LoF variants in 7 genes are present in 0.5% of population. (**C**) GCR of LoF variants in candidate lethal genes. LoF variants in 77 candidate lethal genes are present in 0.5% of population.

Five highest GCRs for each gene set in the general population and in each ethnic group are shown in Figure 3. P/LP and LoF variants in 18 known lethal genes *ABCC6*, *BCKDHB* [MIM *248611], *CEP290*, *DHCR7*, *F7*, *GBA* [MIM *606463], *HBB*, *KIAA0586* [MIM *610178], *KIAA1109* [MIM *611565], *NLRP7* [MIM *609661], *NPC2*, *PKHD1*, *PMM2*, *PYGM* [MIM *608455], *RARS2* [MIM *611524], *SBDS*, *SPEG*, and *SPINK5* [MIM *605010] were present with GCR**≥**0.5% in two or more ethnic groups (Figure 3). Although, each population had unique genes, variants in which are more prevalent in individuals of that ethnicity (Supplementary Figure S2, Table S2). Among P/LP variants, the highest rates were observed for the *DHCR7* (GCR=0.026) and *ASPA* [MIM *608034] (GCR=0.021) genes in Ashkenazi Jewish population, for the *PKHD1* (GCR=0.025) and *GLE1* [MIM *603371] (GCR=0.025) genes in Finnish population, and for the *HBB* gene (GCR=0.025) in South Asians (Figure 3A).

**Figure 3.**
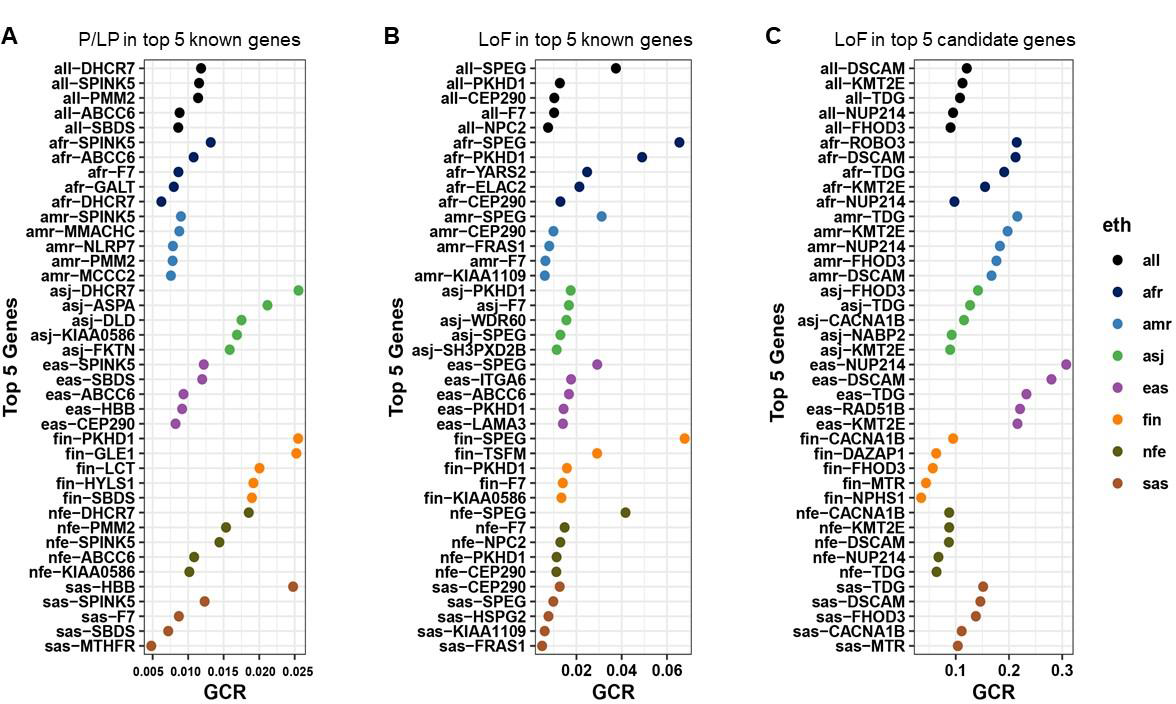
The top five lethal genes with the highest gene carrier rate (GCR) GCR values for the P/LP variants (**A**) and LoF variants (**B**) in the known lethal genes, and (**C**) for LoF variants in the candidate lethal genes. Top five GCR values calculated for all populations (all) are marked in black. Top five genes are also given for 7 ethnic groups African/African American (afr), Latino/Admixed American (amr), Ashkenazi Jewish (asj), East Asian (eas), Finnish (fin), Non-Finnish European (nfe), South Asian (sas). A full list of genes with GCR of 0.005 or higher identified in all populations or any specific ethnic group is given in the Table S2.

Among LoF variants*, SPEG*, a known lethal gene, had the highest GCR in multiple ethnicities (Figure 3B). LoF variants in the *ELAC2* (MIM *605367) (GCR=0.021), *PKHD1*(GCR=0.049), and *YARS2* [MIM *610957] (GCR=0.025) genes were the most frequent in African/African American population, and LoF variants in the *TSFM* gene [MIM *604723] (GCR=0.029) were common in Finnish population (Figure 3B). Table S2 provides a list of 43 known lethal genes that are common (GCR**≥**0.5%) among individuals of a specific ethnicity.

### Gene carrier rates for LoF variants in the “candidate lethal genes”

Top ten genes with the GCR≥0.05 among the human “candidate lethal genes” were *DSCAM* [MIM *602523] (GCR=0.121), *KMT2E* [MIM *608444] (GCR=0.113), *TDG* [MIM *601423] (GCR=0.108), *NUP214* [MIM *114350] (GCR=0.095), *FHOD3* [MIM *609691] (GCR=0.090), *CACNA1B* [MIM *601012] (GCR=0.081), *NABP2* [MIM *612104] (GCR=0.069), *MTR* [MIM *156570] (GCR=0.068), *RAD51B* [MIM *602948] (GCR=0.060), and *ABCF1* [MIM *603429](GCR=0.045) (Table S2). Among the candidate lethal genes, *NUP214*and *DSCAM* genes had the highest GCR for LoF variants in the East Asian ethnic group (Figure 3C) and were also present in the top genes in other populations. Candidate lethal genes with GCR≥0.5% in the general population are listed in the Table S2.

### At-risk couple rates in the general and distinct ethnic populations

At-risk couple rates (ACRs) were calculated for P/LP variants in “known lethal genes”, LoF variants in “known lethal genes”, and LoF variants in “candidate lethal genes”. Cumulative curves for all genes are shown in Figure 4A-C. Up to 0.4% of couples are at risk of having a pregnancy/child affected by one of the autosomal recessive conditions associated with lethal phenotype due to the inheritance of P/LP variants in human “known lethal genes” (Figure 4A). If LoF variants in the known and candidate genes are included, an additional ∼0.2% (Figure 4B) and 9.8% (Figure 4C) of couples in the general population are at risk of adverse reproductive outcomes. Therefore, in the general population at least 10% of couples who are heterozygous for P/LP and LoF variants in lethal genes may be at-risk of a conception with homozygous/compound heterozygous genotypes and a subsequent pregnancy loss. Moreover, at-risk rate depends on the couple’s ethnicity (Figure 4). ACR values were also calculated in each distinct human population. The African/African American and Finnish populations have the highest risk for LoF variants in the known lethal genes” (Figure 4B). Among the “candidate lethal genes”, the highest ACR values is observed for the East Asian ethnicity (Figure 4C).

**Figure 4.**
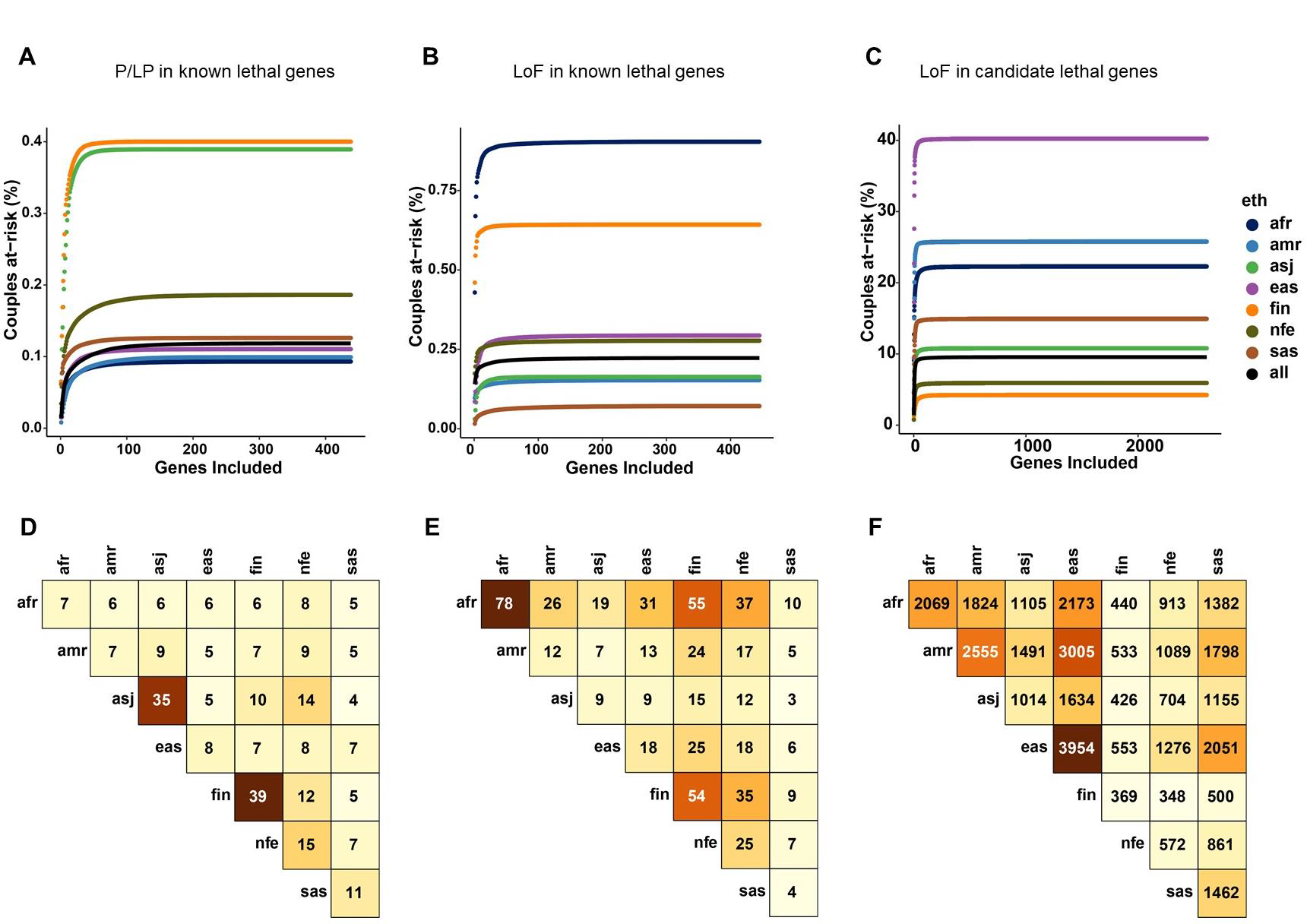
At-risk couple rates (ACRs) Cumulative curves of at-risk couples in distinct human populations for (**A**) P/LP and (**B**) LoF variants in the list of known lethal genes. (**C**) Cumulative curves of at-risk couples for LoF variants in the candidate lethal genes in ethnic groups: African/African American (afr), Latino/Admixed American (amr), Ashkenazi Jewish (asj), East Asian (eas), Finnish (fin), Non-Finnish European (nfe), South Asian (sas). The curve representing the general population (all populations) is highlighted in black. **(D-F)** Intra- and inter-ethnic rate of at-risk couples calculated for 138 genes. ACR values for intra-ancestry couples, when couples are from the same ancestry, and for inter-ancestry couples, when couples are from different ancestries are given as a number of couples at-risk out of 10,000 couples. Number of couples at risk of conceiving an embryo affected by two (**D**) P/LP and (**E**) LoF variants in a gene included in the list of the known lethal genes. (**F**) Number of couples at-risk to have a conception with two inherited LoF variants in a candidate lethal gene.

Risk values were also calculated for the inter-ancestry and intra-ancestry couples for 138 genes with GCR≥0.005 (Figure 4D-F). For P/LP variants, Ashkenazi Jewish and Finnish ethnic groups have the highest risk for intra-ancestry couples (35 and 39 out of 10,000 couples; Figure 4D). For LoF variants in “known lethal genes”, African/African American population had the highest risk for intra-ancestry couples (78/10,000 couples; Figure 4E). In the “candidate lethal genes” the highest risk was calculated for the East Asian ethnic group (3,954/10,000 couples; Figure 4F). Risk values for inter-ancestry couples ranged from 3 to 55 per 10,000 couples for LoF variants in the “known lethal genes” (Figure 4E), and from 348 to 3,005 per 10,000 couples for “candidate lethal genes” (Figure 4F).

We also calculated the probability of an individual to carry P/LP or LoF variants in two or more genes in either “known lethal genes” or “candidate lethal genes”. In the general population, 0.01% (P/LP variants in “known lethal genes”), 0.04% (LoF variants in “known lethal genes”), and 1.4% (LoF variants in “candidate lethal genes)” of individuals are predicted to have variants in two genes from the same gene category (Supplementary Figure S3). Combined, up to 2% of individuals may carry two or more potentially pathogenic variants in known and candidate lethal genes, although the probability to carry variants in multiple target genes may depend on ethnic and familial background of an individual.

### Gene ontology analysis

Using ontology annotations, comparison between known and candidate gene sets was performed. The “known lethal genes” showed enrichment in metabolic processes. Nearly 70% entries were genes which code for proteins classified as metabolite interconversion enzyme (PC00262) or protein modifying enzyme (PC00260) (Figure 5A). In contrast to known lethal genes, candidate lethal genes represent a wide spectrum of proteins (Figure 5B) associated with developmental processes, regulation, multicellular organism processes, as well as metabolic processes. Importantly, deleterious alterations in candidate genes are more likely to disturb early embryo and fetal development, while defects in the known lethal genes are associated with metabolic conditions leading to neonatal and childhood mortality.

**Figure 5.**
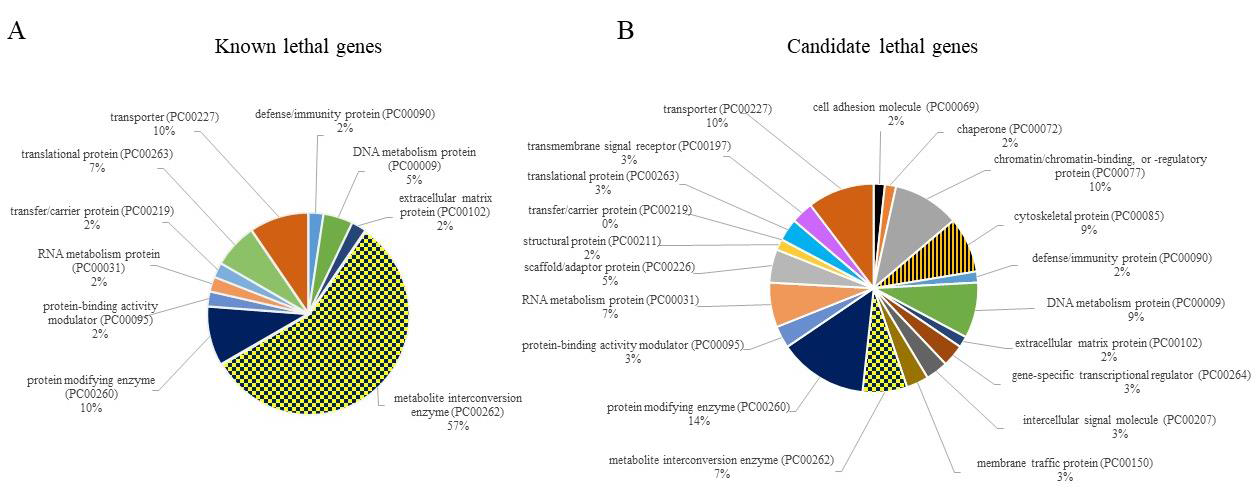
Gene ontology analysis of the known and the candidate lethal genes using PANTHER classification system. (**A**) The main protein class classification of the known lethal genes. (**B**) The main protein class classification of the candidate lethal genes.

## DISCUSSION

In the present study, we performed a population-based analysis and examined allele carrier frequencies of P/LP and LoF variants documented in the gnomAD database for the known and candidate lethal genes. Our study identified 138 genes that may significantly contribute to pregnancy loss due to a high frequency (at least 0.5%) of potentially lethal variants in the general population.

Almost 70% of all human conceptions, both clinically and biochemically detected, are lost.^1,2^ Miscarriages can be traumatic and lead to problems with mental health such as depression, anxiety, and post-traumatic stress disorder.^16,17^ The cause of such losses can be multiple, including environmental,^18^ immunological,^19^ hormonal,^20,21^ anatomic,^22^ and maternal.^1,23,24^ At least 50% of first-trimester miscarriages are not associated with chromosomal abnormalities, suggesting a significant single-gene contribution to euploid losses.^4^ The discovery of lethal genes in humans is hampered by lack of large studies on pregnancy losses and difficulties in evaluation of conceptions at earlier developmental stages. Previous bioinformatic studies have speculated that deleterious variants in ∼600 human genes are associated with perinatal losses, and homozygous LoF alleles for ∼3,400 candidate genes result in non-viable offspring in mice.^11^ Screening for such genes pre-conceptionally, can identify at-risk couples for pregnancy losses, allow preimplantation genetic testing and potentially improve implantation rates for *in vitro* fertilization.

### Contribution of known genes to pregnancy loss and childhood mortality

Only ∼3.7% of known human genes are currently linked to stillbirth and childhood mortality based on reports of neonatal death, fetal demise, and discrepancy between expected and observed birth rates.^11,14,25,26^ Known lethal genes are mainly involved in metabolic processes as revealed by our gene ontology analysis. Pathogenic variants in these genes cause recognizable syndromes, manifestations of which can be viable at birth and *in utero*. Couples who are carriers for pathogenic variants in these genes may experience either livebirth of an affected child or miscarriage, however the factors affecting the severity of the phenotype and onset of fetal/neonatal demise are unknown. In the general population, 5,866 variants in the “known lethal genes” have been classified as P/LP based on their association with specific diseases. Among P/LP, ∼60% are LoF variants. We discovered additional heterozygous 14,532 LoF variants in the “known lethal genes” within gnomAD database (Figure 1). These LoF variants have not been observed in a homozygous state nor linked to a specific disease phenotype in humans, and therefore remain unclassified. Homozygous LoF variants in these genes may cause an identifiable postnatal phenotype or lead to early pregnancy loss. The *DHCR7* gene is one such example among known lethal genes. Pathogenic variants in *DHCR7* result in Smith–Lemli–Opitz syndrome (SLOS [MIM *270400]), an autosomal recessive disorder caused by impaired cholesterol metabolism and characterized by dysmorphic facial features, multiple congenital anomalies involving skeletal, cardiovascular, respiratory, and genitourinary systems. Based on our findings and previously published data,^27,28^ carrier frequency of *DHCR7* P/LP variants ranges from 1/40 to 1/100 individuals among different ethnic groups, with the expected incidence of SLOS between 1/1,600 and 1/10,000 individuals, respectively. The observed incidence of newborns with SLOS in each ethnic group is much lower than expected, suggesting that ∼80% of affected conceptions do not survive to birth^26^. It is likely that compound heterozygous P/LP and LoF conceptions for the *DHCR7* gene are lost before a pregnancy or SLOS phenotype can be recognized. Although *DHCR7* is included in preconception screening, only P/LP variants will be reported. Both partners might be heterozygous for lethal LoF alleles, however their carrier status and the cause of recurrent pregnancy loss may remain undetermined.

Most of the variants are rare, however 1.6% of P/LP variants and 0.37% of LoF variants in 18 “known lethal genes” (*ABCC6, BCKDHB, CEP290, DHCR7, F7, GBA, HBB, KIAA0586, KIAA1109, NLRP7, NPC2, PKHD1, PMM2, PYGM, RARS2*, *SBDS, SPEG,* and *SPINK5)* were seen in 1/200 individuals in the general population. In addition, we identified 43 genes that have P/LP and LoF variants with a frequency of 0.5% or more in ethnic-specific populations (Figure 3A and 3B, Table S2). Deleterious variants in these 61 known genes may greatly contribute to pregnancy losses and neonatal lethality.

### Potential contribution of LoF variants in candidate genes to pregnancy loss

To identify novel autosomal recessive genes, we used bioinformatic and mouse gene candidate approach and focused our analysis on LoF variants in human orthologs. We made significant assumptions considering that like mouse models, the same recessive mode of inheritance will be present in humans. We also assumed that LoF variants in gnomAD database are functional and not polymorphisms. Moreover, it can be argued that we significantly underestimated the carrier rate among candidate genes by focusing on LoF variants, because we did not include missense variants, which pathogenicity at this time is unknown.

Analysis of mouse-lethal human orthologs suggests that 65% (2,385/3,684, Figure 1) of human genes are not linked to any pathology in humans. The remaining, 35% (1,299/3,684) of these genes, are associated with known human diseases, and P/LP variants in 350/1,299 genes (27%) are linked to human lethal phenotypes. Analysis of human genotypes in gnomAD database indicated that homozygous variants in 2,612 genes (Figure 1) were not present in healthy individuals. We identified 77 candidate genes with a high heterozygote frequency of at least 0.5% in general population (Table S2) that may substantially contribute to lethal phenotypes in humans. High rate of heterozygous LoF variants together with the absence of individuals carrying homozygous variants for these genes is consistent with pre/perinatal lethality. Therefore, those genes are likely essential for early human development and, are the candidate genes for autosomal recessive lethal conditions in humans.

Interestingly, among 3,684 genes we did not identify heterozygous LoF variants in 263 human orthologs (Figure 1). Because other types of variants in these genes may still cause fetal lethality and were not included in our analysis, it may underestimate an overall contribution of single gene causes to human pregnancy losses.

In our study, ethnic-specific analysis shows that Finnish, African/African American, and East Asian ethnic groups have a higher rate of conceiving a child with LoF variants in candidate genes (Figure 4A-C). Further clinical evidence should be accumulated to establish association of candidate genes to human lethal phenotypes in different populations.

### Functional differences between known and candidate genes

One of the most important differences between the known genes and candidate genes is their functions and consequently the time point at what they may cause lethality during fetal development. Candidate lethal genes are involved in more global developmental processes (Figure 5). LoF variants in these “essential” genes may not be as tolerable as variants in the known genes, thus, it may cause lethality in earlier stages of embryonic development before any obvious fetal phenotype can be observed. Some of the candidate genes have a very high rate (5-10%) of heterozygous LoF variants in general population (Table S2). For example, LoF variants in *DSCAM* (DS cell adhesion molecule) gene are present in 121 out of 1,000 individuals, and LoF variants in *KMT2E* (lysine methyltransferase 2E) gene are present in 113 out of 1,000 individuals in general population. There are also other genes with common LoF variants in general population such as *CACNA1B* (calcium voltage-gated channel subunit alpha1 B) (81/1,000 individuals), *NUP21* (nucleoporin 214) (95/1,000 individuals), and *TDG* (thymine DNA glycosylase) (108/1,000 individuals). These genes are critical for different global developmental processes such as developing nervous system (*CACNA1B* and *DSCAM*), cellular metabolism, transcriptional regulation, and DNA integrity (*NUP214*, *KMT2E*, and *TDG*).

### Oligogenic causes of pregnancy loss

Since a carrier rate is high for some of the genes, it is possible that both partners will be heterozygous for variants in more than one essential gene. The probability of a couple carrying P/LP or lethal LoF variants in two or more genes is ∼1.5% in the general population (Supplementary Figure S3). This may diminish couple’s probability to conceive an unaffected progeny, as there is a higher chance for each pregnancy to be lost due to multiple monogenic causes or oligogenic etiology.

### Current preconception screenings for the conditions associated with human lethality

Our study provides a set of the relevant and candidate genes connected to human lethality which may be relevant to carrier screening. We identified 138 genes (Table S2) with a high frequency rate of ≥0.5% of damaging variants present in the general population and various ethnic groups. This includes 61 genes with P/LP and LoF variants in a set of known human genes, and 77 candidate genes with LoF variants, seen in all ethnicities. It can be concluded that a small percentage of genes (138/3,080, 4.5%) is responsible for the majority of genetically predisposed non-viable conceptions. Only 29/138 (21%) of the identified genes are currently included in the ACMGG proposed recommendations for preconception genetic carrier screening.^29^ Moreover, utility of preconception carrier screening for patients with pregnancy losses is limited by only reporting P/LP variants. A large fraction of variants incompatible with life will remain unclassified and unlinked to pre/perinatal lethality and pregnancy loss.Accumulation of evidence and further analysis of unclassified variants along with P/LP variants in the setting of lethal phenotypes and reproductive failures are essential to discern variants responsible for fetal lethality and to classify LoF variants based on the ACMGG guideline. Future PGCS panels augmented by genes and variants associated with fetal lethality would be a substantial opportunity to advance public health and reproductive planning.

### Limitations and further directions

We used a set of variants in the known and candidate genes associated with human/mouse lethality based on the existent clinical and experimental evidence from human and mouse studies, as well as gnomAD database to filter for variants that appear to be the subjects of natural selection. This study has focused on autosomal recessive inheritance that can be transmitted from the parents, carriers of at-risk alleles, and potentially identified during preimplantation genetic testing or diagnosed prenatally. We assumed that genes in mouse models of lethality are inherited in autosomal recessive fashion in humans. This assumption is not far-fetched given that 840 out of 1,299 (65%) genes with established phenotype are associated with both, autosomal recessive conditions in humans and lethal phenotype in knockout mice models. We excluded autosomal dominant and X-linked conditions from our considerations to account for hereditary conditions that equally affect conceptions of both sexes. It is possible that the heterozygous variants for some genes are associated with a reduced reproductive fitness or the carrier status itself impacts the ability of an individual to carry pregnancy to term.^30,31^ For example, women with heterozygous variants in the *F2* gene, a coagulation factor II (prothrombin), may develop thrombophilic conditions during pregnancy (RPRGL2 [MIM *614390]), resulting in intrauterine fetal death, irrespectively of a fetus genotype.^31^ Nonetheless, our studies suggest that at least 10% of couples could be at risk of having a conception affected by lethal condition and, therefore, miscarriage. Inclusion of miscarriage-causing genes in the preconception screening panel will significantly augment the detection of at-risk couples as compared to the current carrier screening panels that will identify 0.17-2.52% of couples, depending on their ancestry.^14^

Future studies on large cohorts of diverse patients with pregnancy loss, infertility, fetal demise, stillbirth, and neonatal death need to be conducted to build clinical evidence and confirm an association between the variants in the genes reported here and their effect on fetal and early embryonic development and reproductive outcomes. Identifying multiple unrelated individuals with rare genotypes will be essential to validate the list of potential lethal variants and uncover new gene-phenotype associations. Developing a public database of intolerome (genes and variants that are subjects of biological intolerance and natural selection) will be needed to accumulate genomic findings along with patient’s phenotype, medical history, and reproductive outcomes and will be of great help for future gene discovery and establishment of a precise clinical significance of lethal variants and their penetrance during human development.

## Data Availability

All data produced in the present work are contained in the manuscript

## Acknowledgments

This study was supported by the NIH (R01 HD105256).

## Author Contributions

Conceptualization, A.R., and S.A.Y.; Data curation, J-H.Q., M.A., and S.B.; Formal analysis, M.A., S.B., J-H.Q., and S.A.Y.; Funding acquisition, H.C., E.J., R.B.L., M.S., M.P.S, and A.R.; Writing-original draft, M.A., J-H.Q., and S.A.Y.; Writing-review & editing, M.A., H.C., E.J., R.B.L., M.S., M.P.S, S.A.Y., and A.R.

## Declaration of Interest

The authors declare no competing interests.

## Data and Code Availability

Data are available from the corresponding author.

## SUPPLEMENTARY DATA

### GnomAD variant filtration

The entire set of P/LP and LoF variants was filtered out to exclude: (1) loci with less than 50K confidently measured alleles, (2) alleles that are seen in a heterozygous state in ≥5% of individuals from the general or ethnic population, (3) alleles that are seen in a homozygous state in ≥2, (4) low-quality LoF flag variants, (5) LoF variants flagged as problematic by gnomAD, including sites falling in low complexity (lcr), decoy (decoy) and segmental duplication (segdup) regions, (6) counts from the non-canonical transcripts; (7) duplicated entries.

### GnomAD v2.1.1 ancestral diversity

**Table S1.**
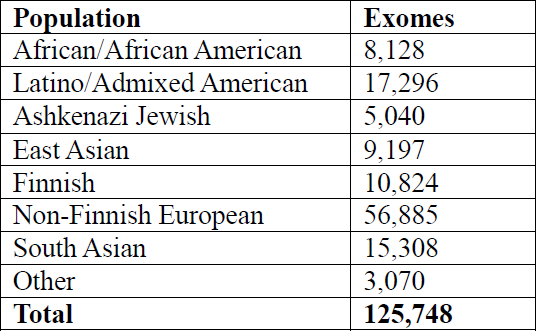
Populations represented in the GnomAD v2.1.1 exome dataset.

### Population risk probability metrics

Variant carrier rate (VCR), frequency of a an individual qualified variant in the general population; gene carrier rate (GCR), a cumulative frequency of all qualified variants in an individual gene; and at-risk couples rate (ACR), the proportion of couples where both partners carry a likely perinatal lethal variant in the same gene were computed as previously described by Guo and Gregg, (doi: 10.1038/s41436-019-0472-7)

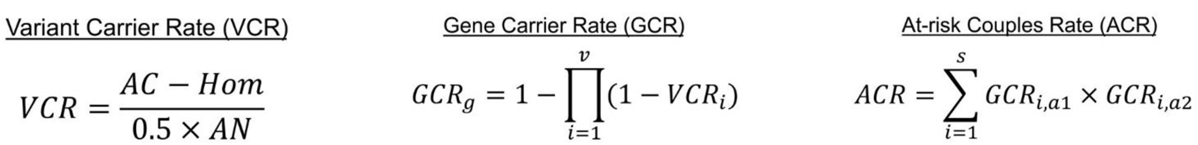

These metrics are given in equations, where AN is the total number of alleles analyzed for the variant, AC is the alternate allele count for the variant, Hom is the number of individuals who are homozygous for the alternate allele; for the GCR of each gene g, the VCRi is the variant carrier rate for variant i, with the cumulative product over v variant targets; for ACR the GCRi,a1 is the GCR for gene i in ancestry-1 and GCRi,a2 is the GCR for gene i in ancestry-2.

**Supplementary Figure S1.**
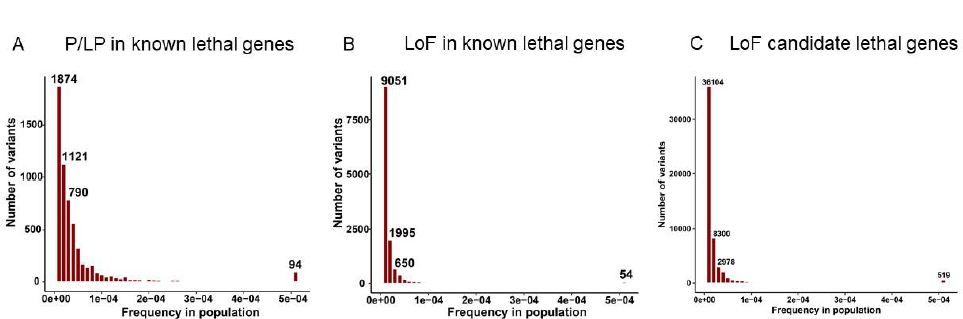
Frequency of qualified variants in the known and candidate lethal genes in the general population. (**A**) VCR of P/LP variants in the known lethal genes. Note, 94 P/LP variants are present with 0.005 allele frequency or seen in 0.5% of population. (**B**) VCR of LoF variants in the known lethal genes. 54 LoF variants are present in 0.5% of population. (**C**) VCR of LoF variants in candidate lethal genes. 519 LoF variants are present in 0.5% of population.

**Supplementary Figure S2.**
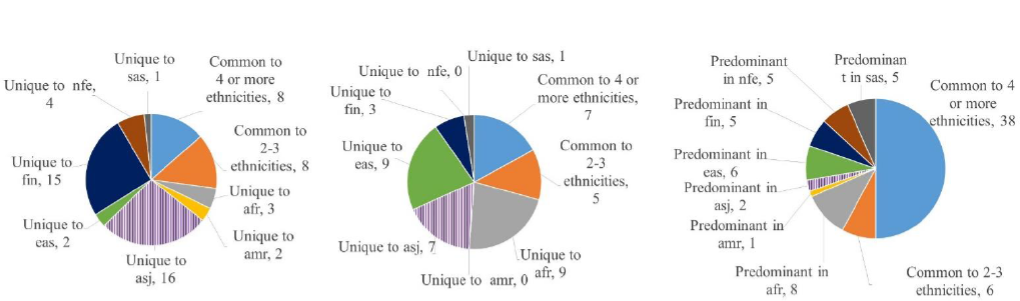
Pan-ethnic and population-biased variants. Common and unique variants in different ethnic groups for (A) P/LP variants in the known lethal genes, (B) LoF variants in the known lethal genes, and (C) LoF variants in candidate lethal genes.

**Supplementary Figure S3.**
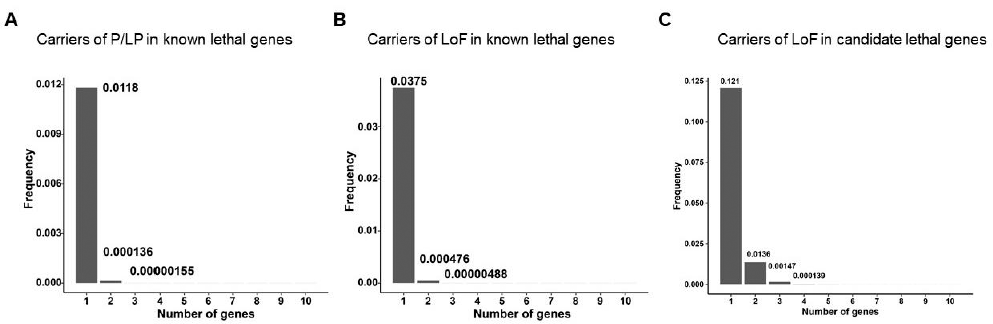
Carrier rates for multiple target genes. Probability for an individual to carry one, two, or several variants when screening for (**A**) P/LP variants or (**B**) LoF variants in the known lethal genes, or for (**C**) LoF variants in the candidate lethal genes.

## Notes

### Competing Interest Statement

The authors have declared no competing interest.

